# Monitoring the first implementation year of the new nutrition labeling regulations in Brazil

**DOI:** 10.1101/2024.04.09.24305563

**Authors:** Camila Aparecida Borges, Carolina Hatsuko Kikuta Batista, Beatriz Silva Nunes, Matheus Nascimento Pureza Castro Leite, Mariana Ribeiro, Laís Amaral Mais, Ana Paula Bortoletto Martins, Ana Clara Duran

## Abstract

**Objective:** This study aimed to monitor the initial 12 months of the implementation of the updated nutrition labeling regulations in Brazil approved in 2020, focusing on the presence and readability of the front-of-package nutrition labeling (FOPNL) on food packages and the presence of added sugars information in the nutrition facts panel.

**Methods:** We used data on nutrition information and FOPNL of 6,829 products launched at Brazilian food retail between November 2022 and October 2023, available at the Mintel – Global New Products Database. We applied eligibility criteria stipulated by regulations to identify products eligible for FOPNL. We classified the foods according to the NOVA classification, identified the products with added sugars information in the nutrition facts panel and those with FOPNL for added sugar, saturated fat or sodium. Moreover, we analyzed the temporal trends in FOPNL presence. Last, a subsample of 202 product labels was analyzed to identify non-compliance with FOPNL readability standards.

**Results:** In the first year of implementation 63.9% of the products analyzed were eligible for at least one critical nutrient’s FOPNL; however, only 12.9% already featured FOPNL by the end of the 12-month implementation period. Among ultra-processed products, 65.1% were supposed to have FOPNL, but only 14.4% did. Less than 30% of sweet cookies, ice cream, tabletop sweeteners, and candies with added sugar in the list of ingredients declared this information in the nutritional facts panel. Analysis of label images revealed non-compliance with FOPNL readability regarding its location on the packaging, FOPNL in removable parts of the packaging or hidden positions, and inadequate color pattern and format.

**Conclusion:** The implementation of the nutrition labeling regulations in Brazil within the first 12 months reached less than 15% of eligible foods and beverages, indicating non-compliance by the food industry. Such inadequacies undermine the expected impact of promoting healthier choices at the point of food purchase.

## INTRODUCTION

When mandatory, front-of-package nutrition labeling (FOPNL) is a practical and effective tool to inform the population about health risks of food products. It also supports purchasing decisions and contributes to preventing non-communicable chronic diseases (NCDs) related to the consumption of ultra-processed products ^1,2^.

In 2020, the Brazilian National Health Surveillance Agency (Anvisa) approved the Resolution of the Collegiate Board (RDC) No. 429/2020 and Normative Instruction (IN) No. 75/2020, which came into effect on October 9, 2022. The new rules improved the readability of the nutrition facts panel, including the mandatory declaration on total and added sugars and the nutrients content by 100g/ml. It also includes mandatory FOPNL on eligible foods and beverages high in saturated fats, added sugars and sodium ^3,4^.

In 2016, Chile was the first country in the world to adopt a warning FOPNL in the format of an octagon in foods high in calories, sugar, sodium and saturated fats. A study evaluating changes in food purchases of the Chilean population after the first phase of the law implementation showed that overall calories purchased declined by 16.4 kcal/per capita/day, also identified declining in sugar, saturated fat, and sodium, when compared with the period before the law ^5^. Another study showed that Chilean families reduced caloric-sweetened beverage purchases by 5.9% and increased purchases of beverages with any non-nutritive sweeteners (NNS) by 26.5% after implementing the warning FOPNL ^6^.

Besides contributing to the reduction of purchasing and consequently the consumption of foods and beverages with high amounts of sugar, fats, and sodium, FOPNL models in general have been seen as a strategy to encourage the food industry to reformulate products, replacing or reducing critical nutrients and ingredients that are risk factors for NCDs ^7^.

In Peru, a study showed a reduction from 9.0 to 5.9 g/100 mL in the median sugar content of beverages, combined with an increase in the use of NNS, leading to a substantial reduction in the number of beverages with a FOPNL warning for this nutrient ^8^.

These Latin American studies emphasize the necessity of ongoing monitoring of the implementation of food labeling policies. The monitoring processes based on data collection in supermarkets have been documented in technical reports by the Chilean Ministry of Health. These reports provide information and standardized procedures to monitor nutrition food labeling laws since 2017, supporting improvement and inspection of the country’s law ^9^. Considering the lack of public data and systems to monitor food labels in Brazil, an alternative to monitoring such regulations is to access data from private companies in the food retail sector. One option is through a database subscription that is frequently updated and contains the nutrition composition and all label information of packaged food and beverages commercialized in the country, such as Mintel – Global New Products Database (Mintel-GNPD) ^10^. Therefore, this study aimed to monitor the initial 12 months of the implementation of the updated nutrition labeling regulations in Brazil approved in 2020, focusing on the presence and readability of the front-of-package nutrition labeling (FOPNL) on food packages and the presence of added sugars information in the nutrition facts panel.

## METHODS

### Sample and presentation of the data source for monitoring

Data were from the Mintel Global New Products Database (GNPD), an online database of newly launched consumer products in global markets. Information on nutrition composition, labeling description, ingredient list and images from food and beverage labels were obtained in this database for this study^10,11^. This database has been widely used in various studies that investigate health claims and/or nutritional content in packaged products ^12–17^. The designation *“newly launched”* includes products that were either introduced to the market or had modifications in formulation or packaging during the study period^11^. Data collection was conducted by trained field researchers (“shoppers”), who photographed product packaging in retail environments and manually entered product information into the Mintel database. The sample comprised products available in food retail establishments located in cities and state capitals from ten Brazilian states: Rio de Janeiro, São Paulo, Paraná, Bahia, Pernambuco, Minas Gerais, Santa Catarina, Goiás, Rio Grande do Sul, Amazonas, and the Federal District (DF).

Mintel-GNPD is a commercial database with restricted access to subscribers, which information available follows attributes recommended by the Pan-American Health Organization (PAHO) ^18^ to analyze public health data: frequency (Mintel has data from food labels since 1996 in Brazil) and timeliness, presents labeling data for the period of implementation of the new labeling rules in Brazil.

All newly launched products available between November 9, 2022, and November 9, 2023 on the Mintel online database, were analyzed in this study, totaling 9,491 products. Despite the new rules implemented on November 9, 2022, the decision was to analyze products included in the database a month later to ensure compliance with the validity period of the regulation, which aligns with the average time of data updating in Mintel-GNPD ^10,11^.

It was possible to analyze a sub-sample of the label images showing all sides of the packaging to identify any inconsistencies in the format of the FOPNL, positioning, size, and color specifications as outlined in IN No. 75/2020.

### Key aspects of RDC No.429/2020 and IN No.75/2020 monitored

The FOPNL is mandatory in all food packages, except for some food categories, if not added with sugar or significant nutritional value from saturated fat or sodium – see more in *IN No.75/2020 (Supplementary Material)*. After the rule came into force in October 2022, three implementation deadlines were established according to the category of packaged products: the first scheduled for October 2023 (for general food products on the market), the second for October 2024 (for products from family farmers or rural entrepreneurs, economic solidarity organizations, individual small businesses, small-scale farmers, artisanal producers, and artisanal foods), and the third for October 2025 (for non-alcoholic beverages in returnable packages). The first deadline was subsequently postponed to October 2024; however, following legal action in support of public health interests, the date was advanced to April 2024 ^19,20^.

However, using the Mintel-GNPD database, it was not possible to identify products from family farmers or similar producers from the second deadline; and no beverages in returnable packaging were available in the study period.

We analyzed 6,829 food and beverages eligible to receive FOPNL according to the food group criteria. A total of 3,112 (32.78%) products that did not meet the eligibility criteria for food groups were excluded (i.e., fruits, vegetables, legumes, root vegetables, cereals, flours, meats, eggs, milk, salt, and others that do not contain added sugars, fats, or sodium, established on IN No. 75/2020 ^4^). Cut-off points stipulated in IN No. 75/2020 were considered to verify the products that needed FOPNL, i.e., added sugar (solid foods: ≥ 15 g/100 g of food; liquids: ≥ 7.5 g/100 g of food), saturated fat (solid foods: ≥ 6 g/100 g of food; liquids: ≥ 3 g/100 g of food), and sodium (solid foods: ≥ 600 mg/100 g of food; liquids: ≥ 300 mg/100 g of food). We also verified the incorporation of total and added sugars declaration on the nutrition facts panel with the quantities per portion, 100 g or 100 mL.

Further evaluation considered the fidelity of the implementation of the regulation regarding requirements for design and legibility of FOPNL, under RDC No. 429/2020^3^, FOPNL declaration must: I – be printed in 100% black on a white background; II – be located on the superior half of the front panel in a unique continuous surface; III – have the same text orientation as the other label information; IV – follow one of the designs defined in Appendix XVII of IN No. 75/2020; V – meet specific requirements of IN No. 75/2020 ^4^, and the FOPNL may not be covered positions or be in removable parts of the package, such as the seal, or located where is difficult to see, such as sealing or twisted areas of the package.

### Study variables

The analysis of macro– and micronutrient data involved standardization, data cleaning—such as the removal of outliers for energy content per 100 g or mL—and adjustment of the variables. Another study compared Mintel-GNPD database and data collected by trained researchers in supermarkets, demonstrating satisfactory agreement ^21^. The study found high consistency (measured by the Intraclass Correlation Coefficient – ICC) for energy value, carbohydrates, total sugars, proteins, total fat, saturated and trans fats, sodium, and fiber. The agreement ranged from good (ICC: 0.6–0.8) to excellent (ICC > 0.8), with no significant differences (p > 0.05) in the mean values of these nutrients per 100 g or mL of product when compared to data collected by trained researchers ^21^.

Dichotomous variables (0=no; 1=yes) were created to classify food that exceeded the established cut-off points by regulation for the three nutrients, and in at least one of the three critical nutrients.

From a variable named “product description”, foods that carried FOPNL were identified through the following searching terms: *“high in added sugar”*, *“high in saturated fat”*, and *“high in sodium”*. This search was conducted in English since it is the language of the Mintel-GNPD. Using content analysis with commands to “string” function from Stata 16.0 statistical package, these terms were identified and converted to dichotomous variables (0=no, 1=yes), named: “received FOPNL for added sugar”, “received FOPNL for saturated fat”, and “received FOPNL for sodium”. A fourth dichotomous variable was named “received at least one FOPNL” when food had at least one and a maximum of three critical nutrients declared on FOPNL.

To aid the monitoring analysis, food and beverages were aggregated by categories and subcategories of most purchased products by Brazilians, as shown by the Household Budget Surveys from 2017/18 ^22^, and by Nova classification of foods based on the extension and purpose of food processing ^23^. However, we did not include the unprocessed or minimally processed foods category (e.g., fruits, vegetables, meats, milk, eggs, cereals, beans, and legumes) as they are foods without the addition of sugar, sodium and fat, so not eligible to receive FOPNL.

### Data analysis

We performed a descriptive analysis of proportion and confidence interval of 95% (CI95%) of food and beverages carrying FOPNL for added sugar, saturated fat, and sodium, as well as the ones eligible considering as parameters the cut-off points of critical nutrients from the nutrient profile model adopted in the IN No 75/2020 and other eligibility criteria by food groups explained above.

Analyses were carried out by categories and subcategories of food groups and by launching classified by Mintel-GNPD, 38.0% referred to new package, 26.0% were new variety or product range extension, 23.0% were new products, 12.0% were relaunching, and 1.0% were new formulation. The proportion of food and beverages eligible and that receiving the FOPNL was calculated for each month of the monitoring period to verify if there was an increase or decrease in regulation compliance regarding the mandatory presence of FOPNL.

The proportion of foods classified as “high in added sugars,” based on the cut-off values established by IN No. 75/2020, was calculated. Among these, the percentage of products reporting added sugars on the nutrition facts panel, typically located on the back or side of the package, was determined. Additionally, the ingredient list of these products was examined to identify the presence of added sugars in the product’s composition. The following ingredients were considered sources of added sugars: cane sugar, beet sugar, sugars from other sources, sucrose, glucose, fructose, lactose, and dextrose, honey, molasses, rapadura, sugarcane juice, malt extract, invert sugar, syrups, maltodextrins, and other hydrolyzed carbohydrates. All analyses were carried out in Stata 16.0.

## RESULTS

During the 12 months, we identified 9,491 food and beverages launched in Brazilian food retail present in the Mintel-GNPD database, from this we analyzed 6,829 that were eligible for FOPL according to the RDC429/2020. According to the NOVA classification, 2.9% of the eligible products were processed culinary ingredients, 18.4% were processed foods, and 78.6% were ultra-processed products. Analyzing the presence of FOPNL by type of launch in the retail sector, from the total of foods with FOPNL, 16.9% were classified as new package, 16.0% new formula, 14.1% re-launch, 10.1% new package, and 8.5% new range of products. Between ultra-processed products, we verified a higher proportion of FOPNL between products classified as new package and new formula.

From the total of products considering eligibles (6,829), around 64% were high in at least one critical nutrient. Among the processed culinary ingredients, 45.3% were high in at least one critical nutrient; among processed foods, 62% were high in at least one critical nutrient; and for ultra-processed products, 65.1% were high in at least one critical nutrient. This proportion was even higher for some subgroups: e.g., between 90.0% and 100.0% of sweet biscuits, margarine, cakes and pies, and chocolates were high in at least one critical nutrient and should present the FOPL (Table 1).

**Table 1.** The proportion (95% CI) of foods and beverages launched in Brazilian retail and considered eligible to receive FOPNL for added sugar, saturated fat, and sodium or at least one critical nutrient during the first year of implementation of RDC N° 429/2020.

| Food Categories | Proportion (95% CI) of eligible products for FOPNL |  |  |  |  |
| --- | --- | --- | --- | --- | --- |
|  | N (%)* | Added Sugar | Saturated Fat | Sodium | At least one critical nutrient |
| **Processed culinary ingredients** | 203 (2.52) | 0.98 (0.24; 3.85) | 27.09 (21.42; 33.61) | 18.22 (13.50; 24.15) | 45.32 (38.60; 52.21) |
| Animal fat | 52 (0.65) | 0 | 100.00 | 65.38 (51.61; 76.98) | 100.00 |
| Starches | 42 (0.52) | 0 | 0 | 0 | 0 |
| Other processed culinary ingredients | 109 (1.35) | 1.83 (0.45; 7.03) | 2.75 (0.10; 8.18) | 2.75 (1.00; 8.18) | 36.69 (28.19; 46.11) |
| **Processed foods** | 1,257 (18.41) | 38.02 (35.38; 40.74) | 36.67 (34.05; 39.37) | 42.88 (40.16; 45.63) | 61.97 (59.25; 64.61) |
| Bread | 102 (1.27) | 48.00 (38.53; 57.68) | 51.96 (42.31; 61.46) | 40.20 (31.14; 49.96) | 50.98 (41.36; 60.53) |
| Cheese | 4 (0.05) | 0 | 75.00 (23.78; 96.64) | 0 | 25.00 (3.35; 76.22) |
| Processed meats | 118 (1.47) | 12.71 (7.81; 20.00) | 28.81 (21.36; 37.61) | 77.97 (69.59; 84.54) | 45.76 (37.00; 54.79) |
| Other processed meats | 1,033 (12.83) | 40.00 (37.12; 43.09) | 35.91 (33.04; 38.89) | 39.30 (36.36; 42.32) | 65.05 (62.09; 67.90) |
| **Ultra-processed products** | 5,369 (78.62) | 64.40 (63.11; 65.68) | 56.79 (55.46; 58.11) | 51.31 (49.97; 52.64) | 65.06 (63.78; 66.32) |
| Sausages and other reconstituted meat products | 299 (3.71) | 76.25 (71.11; 80.73) | 21.07 (16.81; 26.06) | 94.64 (91.44; 96.69) | 76.92 (71.80; 81.35) |
| Sweet biscuits | 442 (5.49) | 92.53 (89.68; 94.64) | 93.89 (91.23; 95.77) | 79.63 (75.62; 83.14) | 94.11 (91.50; 95.96) |
| Savory biscuits | 490 (6.09) | 59.39 (54.97; 63.65) | 71.02 (66.84; 74.87) | 70.00 (65.79; 73.89) | 73.26 (69.17; 76.99) |
| Margarine | 19 (0.24) | 10.53 (2.64; 33.74) | 100.00 | 94.74 (70.60; 99.26) | 100.00 |
| Cakes and pies | 335 (4.16) | 91.64 (88.15; 94.17) | 84.17 (79.86; 87.71) | 42.98 (37.78; 48.35) | 97.61 (95.29; 98.80) |
| Sliced bread | 311 (3.86) | 67.20 (61.78; 72.19) | 78.13 (73.20; 82.38) | 41.47 (36.12; 47.04) | 44.05 (38.62; 49.62) |
| Candies in general | 633 (7.86) | 72.19 (68.57; 75.54) | 37.75 (34.06; 41.60) | 30.96 (27.48; 34.67) | 79.30 (75.97; 82.28) |
| Carbonated sweetened beverages | 172 (2.14) | 69.76 (62.49; 76.16) | 0 | 73.25 (66.14; 79.33) | 27.32 (20.18; 34.46) |
| Chocolate | 398 (4.94) | 95.72 (93.23; 97.34) | 97.73 (95.71; 98.81) | 12.31 (9.43; 15.91) | 99.25 (97.68; 99.75) |
| Noodles | 108 (1.34) | 10.18 (5.72; 17.46) | 35.18 (26.77; 44.62) | 29.63 (21.79; 38.89) | 18.51 (12.26; 26.97) |
| Ready-to-eat meals | 333 (4.14) | 48.64 (43.31; 54.01) | 79.27 (74.58; 83.29) | 81.68 (77.15; 85.47) | 49.24 (43.90; 54.60) |
| Non-carbonated sweetened beverages | 539 (6.69) | 41.37 (37.28; 45.58) | 15.21 (12.42; 18.50) | 31.35 (27.57; 35.39) | 28.57 (24.91; 32.53) |
| Dairy beverages | 262 (3.25) | 32.82 (27.40; 38.74) | 23.66 (18.90; 29.18) | 39.69 (33.94; 45.74) | 7.63 (4.97; 11.53) |
| Ice cream | 214 (2.66) | 73.83 (67.53; 79.28) | 77.57 (71.49; 82.66) | 41.59 (35.17; 48.30) | 49.53 (42.88; 56.20) |
| Sauces and condiments | 458 (5.69) | 49.34 (44.78; 53.91) | 44.76 (40.26; 49.34) | 64.19 (59.69; 68.45) | 72.49 (68.22; 76.38) |
| Tabletop sweetener | 17 (0.21) | 35.29 (16.78; 59.59) | 0 | 35.29 (16.78; 59.60) | 29.41 (12.80; 54.19) |
| Others UPPs | 339 (4.21) | 53.68 (48.35; 58.93) | 69.03 (63.90; 73.72) | 43.95 (38.75; 49.28) | 76.69 (71.89; 80.89) |
| **TOTAL** | **6,829 (100.00)** | **57.66 (56.49; 58.83)** | **52.20 (51.02; 53.38)** | **48.77 (47.59; 49.96)** | **63.90 (62.75; 65.03)** |

CI 95%: 95% Confidence Interval; FOPNL: Front-of-package Nutrition Labeling; \*total number of foods that were considered eligible to apply the cut-off points for the critical nutrients studied.

In this first year of implementation, only 12.9% of the eligible food and beverages comply with the regulation. Only 5.4% of processed culinary ingredients, 7.9% of processed foods, and 14.4% of ultra-processed products received a FOPNL for at least one of the three critical nutrients. From the analyses, more than 60.0% of ultra-processed should have FOPNL. Among this subgroup, chocolates and margarine are the products that had more FOPNL for added sugar, saturated fat and sodium, with around 40.0% of the products complying with the regulation (Table 2).

**Table 2.**
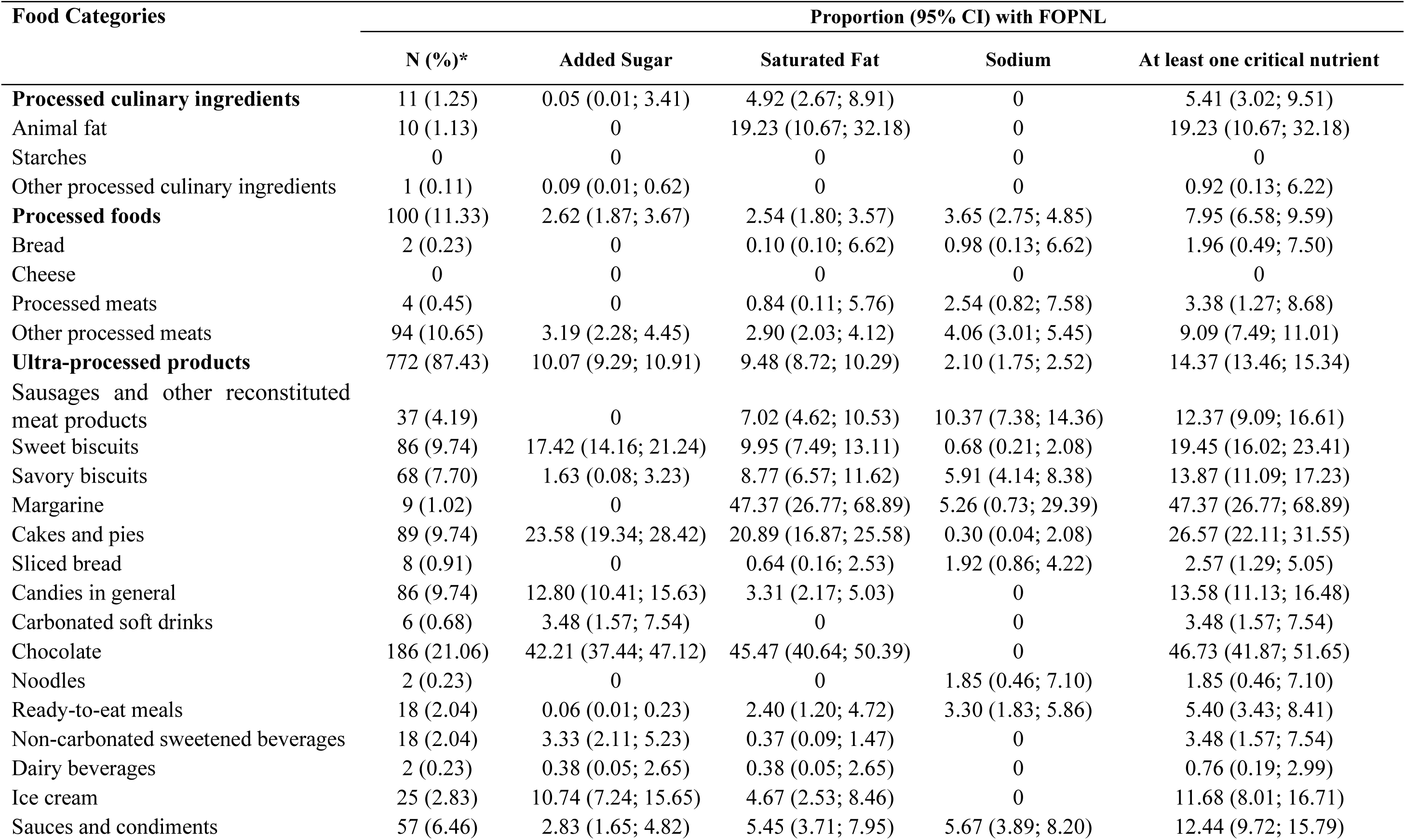
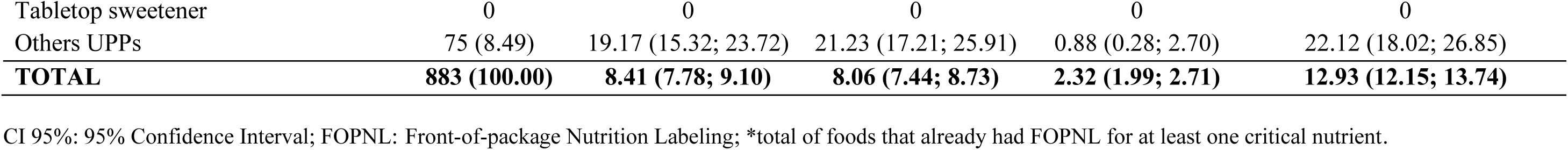
The proportion (95% CI) of foods and beverages launched in Brazilian food retail with FOPNL for added sugar, saturated fat, and sodium or at least one critical nutrient during the first year of implementation of RDC N° 429/2020.

Only 20.3% of the processed foods high in this nutrient have declared the information of added sugars, while for ultra-processed products, only half of the products high in added sugar had declared it. In some subcategories of ultra-processed products, such as sweet biscuits, ice creams, tabletop sweeteners, candies, sausages, and other reconstituted meat products, the proportion of added sugar declaration on the nutrition facts panel was below 30.0%, and all of them had added sugar in the ingredient list (Table 3).

**Table 3.** The proportion of foods and beverages high in added sugar and declared added sugar information in the nutritional facts panel.

| **Food Categories** | **n (%)** | **High in added sugar** | **Added sugar on nutrition facts panel\*** |
| --- | --- | --- | --- |
| **Processed culinary ingredients** | 203 (2.52) | 0.98 (0.24; 3.85) | 0 |
| Animal fat | 52 (0.65) | 0 | 0 |
| Starches | 42 (0.52) | 0 | 0 |
| Other processed culinary ingredients | 109 (1.35) | 1.83 (0.45;7.03) | 0 |
| **Processed foods** | 1,257 (18.41) | 38.02 (35.38; 40.74) | 20.29 (16.92; 24.14) |
| Bread | 102 (1.27) | 48.00 (38.53; 57.68) | 16.32 (8.37; 29.39) |
| Cheese | 4 (0.05) | 0 | 0 |
| Processed meats | 118 (1.47) | 12.71 (7.81; 20.00) | 13.33 (3.35; 40.55) |
| Other processed meats | 1,033 (12.83) | 40.00 (37.12; 43.09) | 21.01 (17.35; 25.21) |
| **Ultra-processed products** | 5,369 (78.62) | 64.40 (63.11; 65.68) | 33.54 (31.99; 35.13) |
| Sausages and other reconstituted meat products | 299 (3.71) | 76.25 (71.11; 80.73) | 21.49 (16.63; 27.29) |
| Sweet biscuits | 442 (5.49) | 92.53 (89.68; 94.64) | 28.36 (24.20; 32.92) |
| Savory biscuits | 490 (6.09) | 59.9 (54.97; 63.65) | 31.27 (26.20; 36.83) |
| Margarine | 19 (0.24) | 10.53 (2.64; 33.74) | 50.00 (5.88; 94.11) |
| Cakes and pies | 335 (4.16) | 91.64 (88.15; 94.17) | 34.20 (29.11; 39.68) |
| Sliced bread | 311 (3.86) | 67.20 (61.78; 72.19) | 48.32 (41.62; 55.09) |
| Candies in general | 633 (7.86) | 72.19 (68.57; 75.54) | 28.88 (24.91; 33.21) |
| Carbonated soft drinks | 172 (2.14) | 69.76 (62.49; 76.16) | 37.50 (29.31; 46.47) |
| Chocolate | 398 (4.94) | 95.72 (93.23; 97.34) | 55.11 (50.08; 60.04) |
| Noodles | 108 (1.34) | 10.18 (5.72; 17.46) | 0 |
| Ready-to-eat meals | 333 (4.14) | 48.64 (43.31; 54.01) | 31.48 (24.80; 39.03) |
| Non-carbonated sweetened beverages | 539 (6.69) | 41.37 (37.28; 45.58) | 26.90 (21.49; 33.11) |
| Dairy beverages | 262 (3.25) | 32.82 (27.40; 38.74) | 38.37 (28.73; 49.02) |
| Ice cream | 214 (2.66) | 73.83 (67.53; 79.28) | 18.35 (13.06; 25.17) |
| Sauces and condiments | 458 (5.69) | 49.34 (44.78; 53.91) | 25.66 (20.38; 31.75) |
| Tabletop sweetener | 17 (0.21) | 35.29 (16.78; 59.59) | 16.66 (2.28; 63.14) |
| Others UPPs | 339 (4.21) | 53.68 (48.35; 58.93) | 42.85 (35.86; 50.15) |
| **Total** | **6,829 (100.00)** | **57.66 (56.49; 58.83)** | **31.91 (30.48; 33.39)** |

\*Proportion calculated only when the product was considered high in sugar according to the criteria of RDC no. 429/2020.

Figure 1 shows the monthly evolution of foods that received FOPNL for at least one critical nutrient in the first 12 months of RDC No 429/2020 implementation and the proportion of foods eligible to receive FOPNL launched in the Brazilian market. At the beginning, the proportion of foods that had received FOPNL was around 10%, and in the end, this proportion increased to 30%. On one hand, we verified that the proportion of eligible foods decreased gradually over the months. On the other hand, the proportion was sustained at 60.0% and 70.0%, almost the same amount as at the beginning of the monitoring, which may indicate the non-compliance of FOPNL incorporation by the food industries of the country.

**Figure 1.**
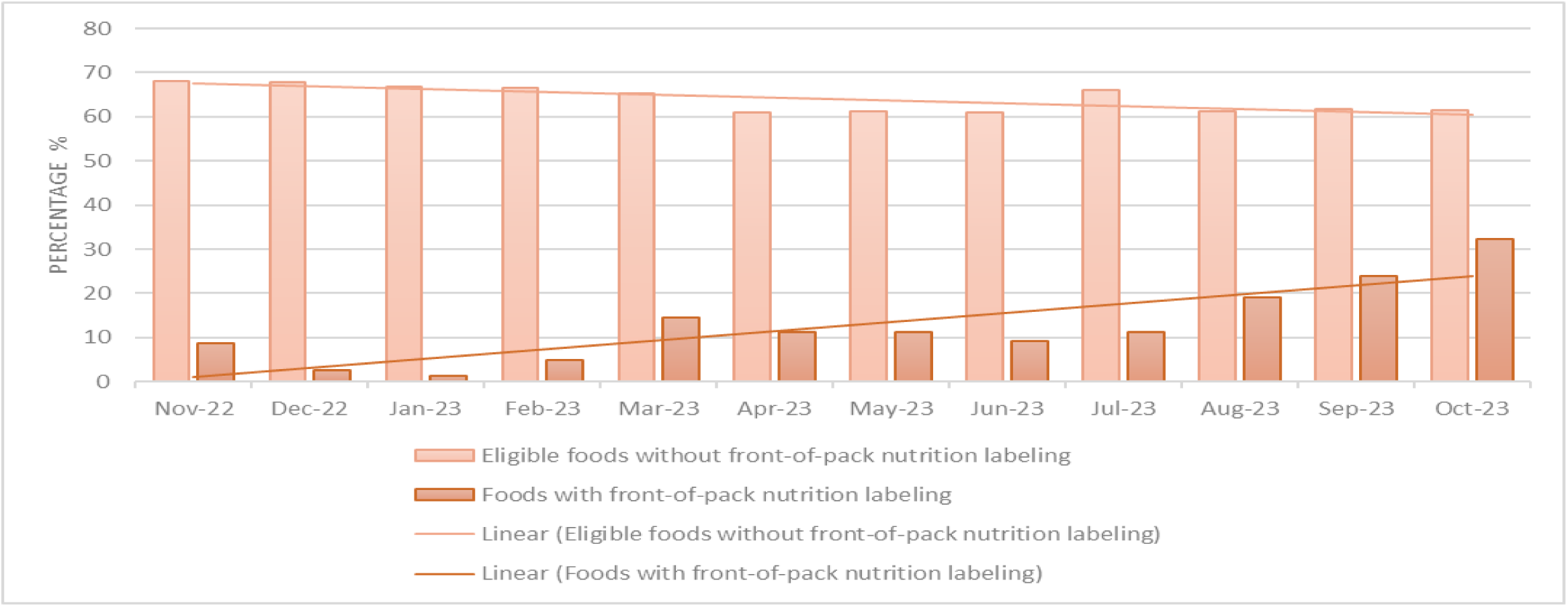
Evolution of the prevalence of front-of-pack nutritional labeling implementation and eligible products over the months during the monitoring period.

Also, inadequacies in FOPNL legibility were observed in a sub-sample of 202 label images launched in the first six months of monitoring. From this total, 61.1% were chocolates, 17.8% baked goods, 6.7% snacks, 4.3% sweet fillings for bread, 4.3% were sauces and seasoning, 2.4% were dairy products. Around 41% of the products had the magnifying glass positioned on the side of the package and not in the frontal face aligned with the other information. In 12.0% of the products, the FOPNL was in removable or hidden parts of the package, 1.0% in the back of the package, and 0.05%, the color pattern and format were different from recommended (Table 4).

**Table 4.**
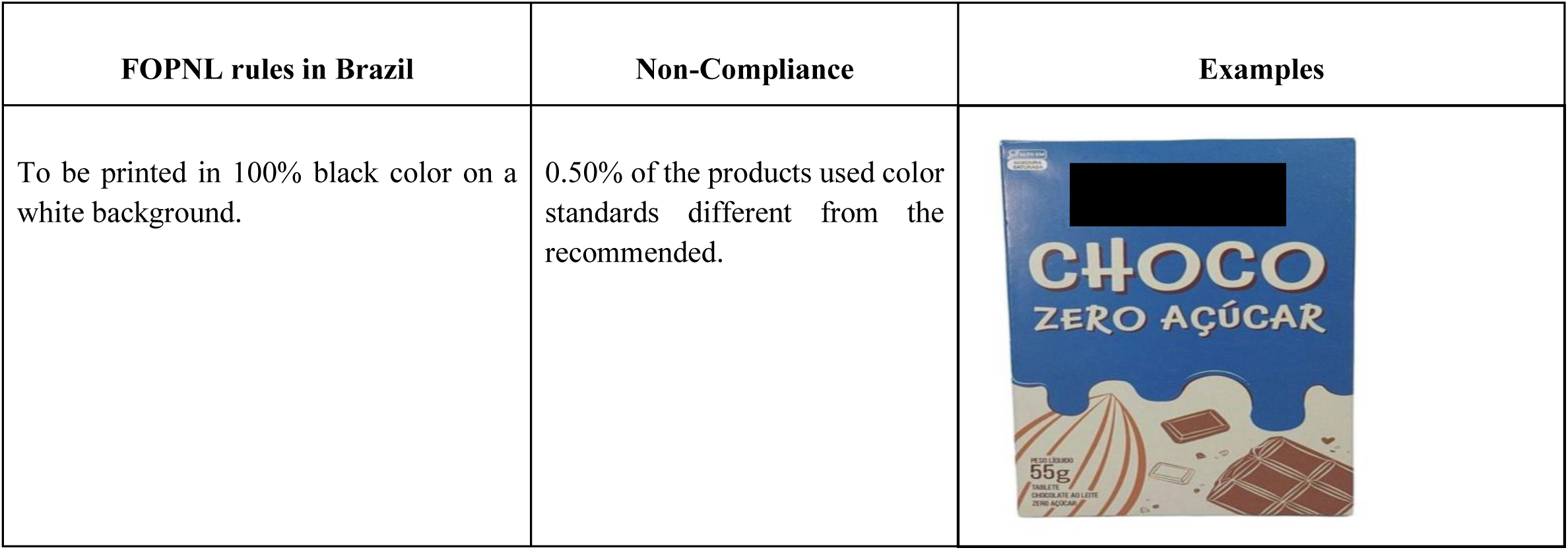
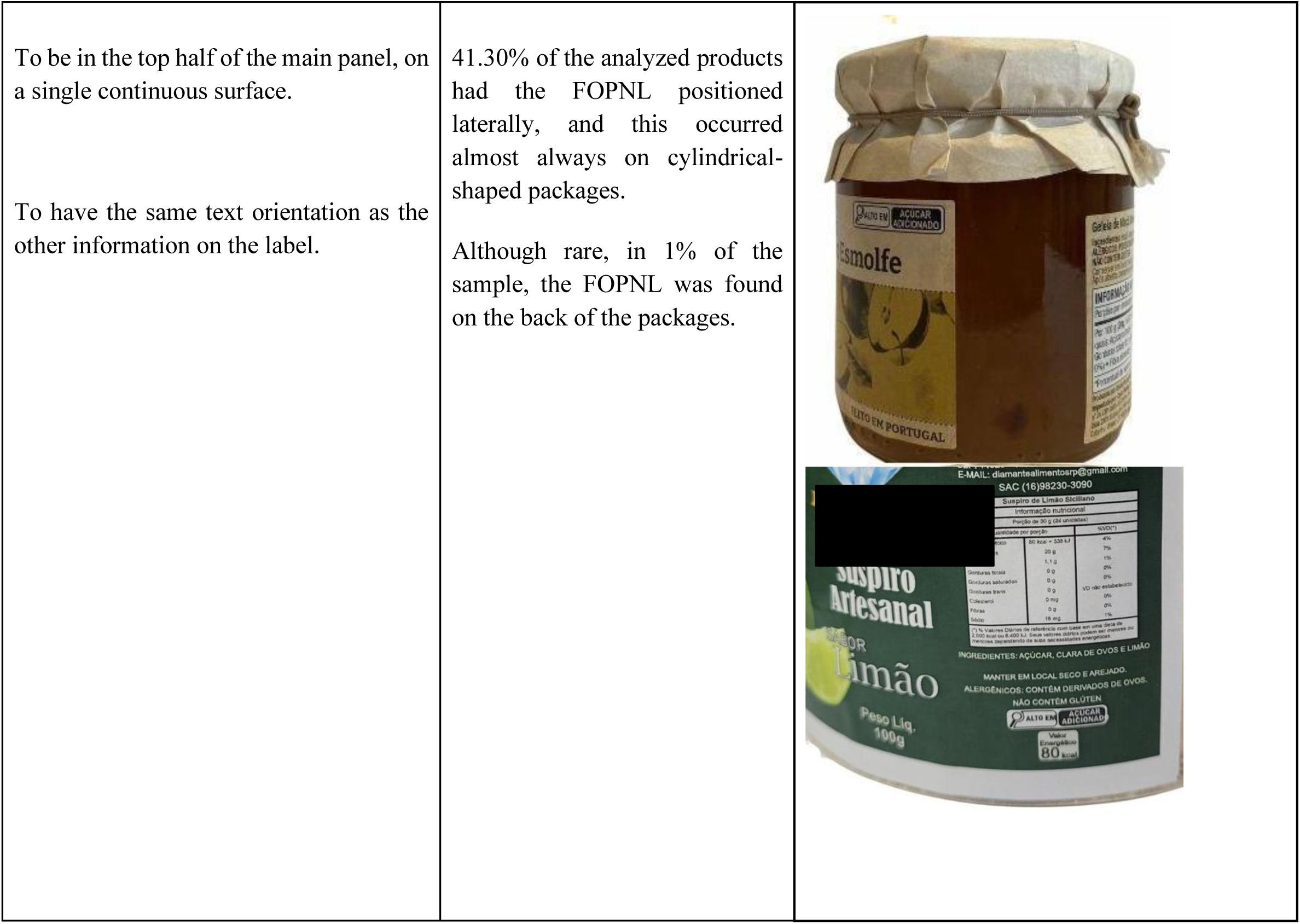
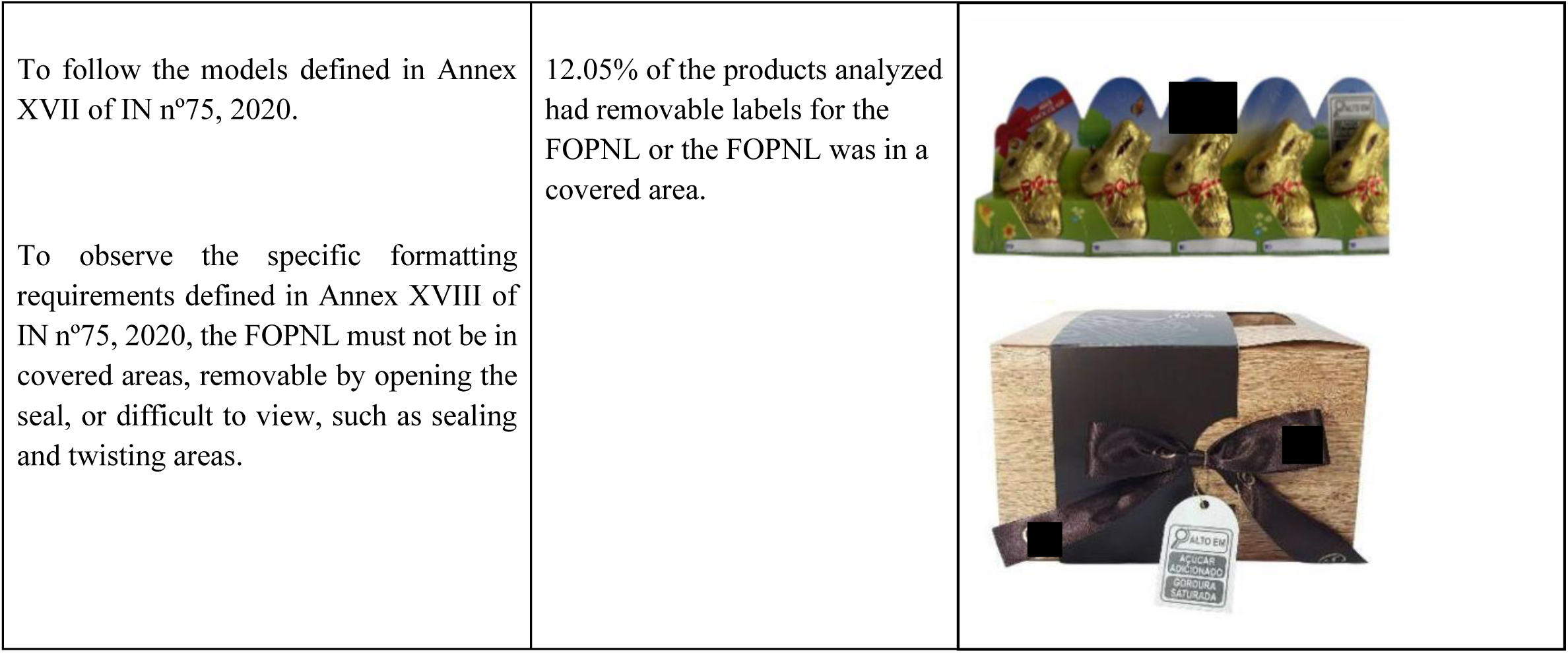
Characterization and illustrations of non-compliance in Brazilian products with FOPNL from 11/09/2022 to 06/09/2023.

## DISCUSSION

This study briefly showed that, of all the foods analyzed, around 63.9% were eligible to receive the FOPNL for at least one nutrient. Yet only 12.9% (or 5 times less) already had this information on the label. The situation is even worse among ultra-processed products, where 65.1% should receive the FOPNL, and just 14.4% were complying with the regulation in the first year of implementation. In addition, products such as sweet biscuits, ice cream, tabletop sweeteners, candies in general, sausages, and other reconstituted meat products high in sugar had a low proportion of added sugar information in the nutrition facts panel (<30.0%). After analyzing the label images of a sample of products in the first six months, inadequacies were observed in the standardization and legibility of the FOPNL on food packages.

In Brazil, another study collected primary data from packaging directly in supermarkets from May to October 2023, adding up to 2,145 products found in five Brazilian states and focusing on characterizing the products with the presence of FOPNL ^24^. Herein, we used a commercial database with a sample of 6,829 products launched in the retail sector over 12 months, with the additional aim of verifying eligibility and inadequacies in the FOPNL readability. However, similarities among the studies can be observed where both showed that, among the products with a FOPNL, the majority were high in added sugar and/or saturated fat and most of them classified as ultra processed foods.

In Latin America, countries that have a mandatory implementation of warning FOPNL, such as Chile and Uruguay (octagon format), have presented monitoring analyses and technical reports from governments following the compliance process by food industries and consumers’ understanding of the presence of labeling on food in the first months of implementation ^9,25^. In Uruguay, a study with consumers in the first month of the law’s implementation showed a high level of awareness and self-reported use of the FOPNL ^25^.

In addition, a comparison between before and after implementation showed that the FOPNL increased consumers’ ability to use nutrition information to compare products and identify products with excessive sugar, fat, saturated fat and sodium content ^25^. In Chile, the governmental technical report of 2017, one year after the law implementation, including mandatory warning FOPNL and restriction of child-directed marketing, showed that 71% of food retailers were complying with the law ^9^.

Our study showed that in the first 12 months of implementation of RDC No. 429/2020 ^3^, only 13% of new products launched in retail stores had FOPNL for at least one critical nutrient. A monitoring approach carried out in Chile in the first year of implementation of mandatory front-of-package nutrition labeling showed that around 60% of products already had FOPNL ^26^, which shows that Brazil may be implementing at a slower pace compared to other Latin American countries. One hypothesis for the slow pace may be different implementation deadlines for different types of products, for example, family agribusiness and returnable packaging, whose implementation deadlines were April 2024 and October 2025, respectively, and/or industry interference in the process, such as the proposal of RDC No. 819/2023 ^19^.

In Brazil, adopting a new labeling regulation with FOPNL took a long time and was strongly influenced by the food industry at various stages of the regulatory process. This influence has yielded more delays ^27^, including the publication of RDC No. 819/2023 ^19^, which determined that the food industry could use old packaging in storage without incorporating the FOPNL for longer than determined by RDC No. 429/2020 ^3^. The industry’s interference in delaying the regulatory process is also reflected in the large number of foods we identified as high in that did not present the FOPNL for critical nutrients.

It is widely known that ultra-processed products most often exceed the amounts of sugar, fat and sodium ^21^ and have been associated with a higher risk of obesity, cancer, and cardiovascular diseases, among other diseases ^28^. The consumption of these foods is associated with poorer diet quality ^29^ and environmental impact ^30^. Therefore, as soon as all ultra-processed products in Brazil that exceed the thresholds for critical nutrients are complying with the regulations, the sooner consumers will improve their food choices and, consequently, reduce the prevalence of obesity and other NCDs in the country.

Our study showed that 64.4% of ultra-processed foods had high levels of added sugars according to the nutrient profile model adopted in Brazilian nutritional labeling regulations. Of these, 10.7% had a magnifying glass on the front of the package warning about the presence of excessive added sugar, and 33.5% contained information about added sugar in the nutrition facts panel. This discrepancy in the inclusion of complete information on added sugar on the labels of ultra-processed products could illustrate difficulties in the process of implementing RDC429/2020 by food industries in the country. On January 19, 2023, Anvisa published a second edition of a document with questions and answers to support companies in implementing the new labeling standards ^31^. This document provides information on the possible sources of added sugars and could help various industries, especially smaller ones, to comply with health legislation. However, the document was published several months after the regulation began to be implemented in the country.

Sweetened beverages have been associated with diabetes, obesity, and cardiovascular disease ^32^. They have been the subject of public policy discussions on taxation and warning FOPNL to reduce consumption by the population ^33^. In Chile, the volume of beverage purchases that were high in added sugar decreased by 22.8 mL/per capita/day when compared to the scenario before the law implementation ^34^. Our study found that only 3.5% of carbonated soft drinks had FOPNL for high added sugar, yet almost 70% of these products were eligible to comply with the regulation.

The World Health Organization (WHO), the United Nations Children’s Fund (UNICEF), and the World Heart Federation (WHF) recognize the role that food labeling policies, including FOPNL, play in promoting healthier choices and preventing NCDs related with the consumption of unhealthy foods ^35–37^. When mandatory, FOPNL leads the reformulation of food industry products ^7,8^, aiming to reduce or replace critical nutrients, such as replacing sugar with sweeteners ^8^. However, these replacements are not considered the best alternatives, as the products remain being ultra-processed and there are other health risks related to the ingredient’s substitution.

According to a recent technical report published by the WHO, there are potential undesirable effects resulting from the prolonged use of sweeteners, such as an increased risk of type 2 diabetes, cardiovascular disease, and mortality in adults^38^. In this context, constant monitoring of the nutrition composition of foods and beverages to identify reformulation processes is crucial to guarantee the nutritional quality of reformulated foods and to verify the possible health impacts that these reformulations could bring to the population.

In Brazil, there is no data showing changes in the nutrition profile of products from reformulation or changes in the food purchases due to the implementation of the updated nutrition food labeling legislation yet. From a public health perspective, monitoring nutrition composition and food labeling of packaged food and beverages provides information about the healthiness of the consumer’s food environment. It also supports the planning, implementation, and improvement of interventions in public health ^39^.

There are different ways to monitor this information, such as laboratory analyses (which are costly and tricky to develop on a large scale), data collection in food stores (requires team recruitment, training, data treatment and typing to create a database), web-scraping of available data in online sources, and data from label images shared by consumers using smartphones applications (e.g. *Desrotulando* App)^39^. However, few strategies can capture the dynamic changes in the food market ^40^.

One of the strengths of our study was the use of validated commercial data ^21^, which provided a monthly updated database with a large sample of foods and beverages launched in Brazilian food retail. These data made it possible to carry out the monitoring analysis quickly and with country level representation. Among the limitations about the food and beverage dataset used is that these products are characterized by some type of launch, and in this case, products available in food retail that did not fit these characteristics were not collected by Mintel-GNPD. Therefore, only an audit process at supermarkets would provide better estimates of the presence of FOPNL in the food available in food retails, making it possible to analyze those products that were already consolidated in the market and were not launched. However, given the lack of data on the labeling of packaged foods and beverages that could be used to monitor the implementation of RDC No. 429/2020 in the country, we consider the information in the Mintel-GNPD database to be extremely relevant in this initial process, and with that, researchers could support public actors with scientific evidence.

In conclusion, the implementation of RDC No. 429/2020 was slow and gradual in the initial 12 months, highlighting the hardships faced by food industries to meet the regulatory deadlines. Besides, inadequacies regarding FOPNL legibility may compromise consumers’ healthier choices when purchasing food.

## STATEMENTS

### Data availability statement

All data produced in the present work are contained in the manuscript

### Ethics approval statement

Not apply

### Permission to reproduce material from other sources

Not apply

